# CGM accuracy and reliability compared to point of care testing in older inpatients with comorbid type 2 diabetes and cognitive impairment

**DOI:** 10.64898/2026.03.27.26349485

**Authors:** Busra Donat Ergin, Katharina Mattishent, Anne-Marie Minihane, Richard IG Holt, Helen R. Murphy, Ketan Dhatariya, Michael Hornberger

## Abstract

**Background:** Older in-patients have a higher prevalence of diabetes and cognitive impairment compared to those living without diabetes or cognitive decline. Cognitive impairment can complicate glucose management more challenging, as patients may forget to measure their blood glucose or report symptoms. Investigating the accuracy of continuous glucose monitoring (CGM) compared to usual care will inform clinical decision-making in this vulnerable population.

**Aim:** To compare CGM derived glucose metrics and point-of-care tests (POCT) in older in-patients with type 2 diabetes and cognitive impairment, and to investigate the analytical and clinical accuracy of CGM in the hospital setting.

**Methods:** Thirty-two older people with comorbid type 2 diabetes and cognitive impairment were recruited within a UK tertiary care hospital. All participants were CGM-naive and wore blinded Dexcom G7 sensors for up to 10 days, while receiving. Usual care during their hospital stay including POCT. Key accuracy metrics comprised the mean absolute relative difference (MARD), median absolute relative difference (median ARD), Clarke Error Grid (CEG) and, correlation (R^2^). The proportion of CGM readings within ±20% of reference glucose values when the reference was >5.6 mmol/L, or within ±1.1 mmol/L when ≤5.6 mmol/L (±20%/1.1 mmol/L) was calculated to assess analytical and clinical accuracy.

**Results:** Thirty participants completed the study. CGM-derived mean glucose for time in range (TIR, 4-10 mmol/mol) was 36.2% (range 0-90%), time above range (TAR ≥10 mmol/mol) was 62.8% and time below range (TBR ≤3.9 mmol/mol) was 1%. Mean POCT-based TIR was 41%, TAR was 57.2% and TBR 2%, showing broadly comparable glucose metrics. CGM accuracy analysis showed a MARD of 17.4%, median ARD of 12.2% and the outcome of ±20%/1.1 mmol/L was agreement of 72%. CEG analysis revealed that 99.3% of paired readings fell within the clinically acceptable Zones A and B, with a strong correlation between CGM and POCT (R^2^=0.82). CGM identified more hypoglycaemic readings, particularly at night.

**Conclusion:** CGM and POCT produced largely similar glucose metrics in older in-patients with diabetes and cognitive impairment. CGM demonstrated clinically acceptable accuracy, and captured additional hypoglycaemia, undetected by routine POCT. These findings suggest that CGM may be a viable and potentially advantageous alternative to POCT in this high-risk population.

## Introduction

Type 2 diabetes (T2DM) and dementia are common, progressive long-term conditions associated with ageing that predominantly affect older adults [1], [2], [3]. People with diabetes are approximately twice as likely to develop all-cause dementia compared with those without diabetes and typically experience an earlier onset of cognitive decline [2], [3].

Comorbid T2DM and dementia is highly prevalent in hospital populations [4], [5]. Approximately 25% of inpatients in acute hospital beds have dementia, with the prevalence of individuals with all-cause dementia and T2DM estimated at 8.3% [4]. Moreover, the comorbidity is associated with having longer hospital stays than those with either T2DM or dementia [6], and comorbid patients have also higher hospital mortality and readmission rates [7]. This is due to several factors particularly the higher incidence of dysglycaemia (hypo- and hyperglycaemia) in people comorbid T2DM and dementia [6]. Asymptomatic hypoglycaemia, particularly, emerges as a significant acute complication of diabetes, especially in people with cognitive impairment [8].

Both hyperglycaemia and hypoglycaemia are associated with harm in a general inpatient population [9]. Given the increasing incidence of comorbid dementia and T2DM in elder people, it seems essential to consider better glycaemic management in this vulnerable population with T2DM and cognitive impairment. Retrospective data has shown that glycaemic management is the key preventable factor to reduce acute complications of diabetes, and it may be optimised with the use of continuous glucose monitoring (CGM) systems in older people. CGM is associated with reduced hypoglycaemia risk in inpatients and real-time CGM detects asymptomatic dysglycaemia with its frequent and continuous interstitial glucose measurements [10], [11], [12], [13].

However, evidence on CGM accuracy in hospitalised older adults with comorbid type 2 diabetes and cognitive decline, to our knowledge, does not exist. Previous studies explored clinical accuracy and reliability of different CGM sensors compared to point-of-care testing (POCT) in T1DM or T2DM without cognitive impairment/dementia [13], [14], [15], [16], [17]. Further evidence is needed regarding the clinical accuracy and reliability of next-generation CGM systems, compared with POCT particularly in individuals with comorbid T2DM and cognitive impairment.

The current study rectifies this shortcoming by reporting the clinical accuracy and reliability of CGM compared with POCT in hospitalised older adults with comorbid T2DM and cognitive impairment. The analyses are based on a blinded feasibility study [18]. Due to the blinded nature, neither participants nor clinician had an insight into the CGM readings, providing a more objective approach towards comparing CGM and POCT in this vulnerable population. We hypothesised that CGM and POCT would largely align, similar to previous findings in T2DM alone; however, CGM might be able to pick up hypoglycaemic changes earlier than POCT due to its continuous measurements.

### Methodology

A single-centre, prospective, observational study in older in-patients with T2DM and cognitive impairment on non-intensive care units (non-ICU) at the Norfolk and Norwich University Hospital, UK.

Eligible participants were inpatients, older (≥65 years) adults with T2DM for at least 6 months and on diabetes management medications (i.e. not managed by diet/lifestyle alone). Participants were also required to have an Abbreviated Mini-Mental Test (AMT) score equal or less than 8 (out of 10) and/ or a Mini-Addenbrooke’s Cognitive Examination (Mini-ACE) UK Version A score ≤21 (out of 30), indicating cognitive impairment. The AMT is a commonly used brief cognitive screening tool within hospitals in the UK. The Mini-ACE is a validated cognitive screening tool sensitive to earlier cognitive changes using two cut-offs (25 or 21 out of 30, and we used 21 cut off for this study) [19].

Potential participants were excluded if they did not have mental capacity and/or were on end-of-life care, as confirmed by the clinical care teams. We also excluded people who were current CGM users, were undergoing dialysis treatment and showed evidence of skin irritations on the upper arm or abdomen where the CGM sensor would potentially be fitted. We also excluded people who had other major neurological or psychiatric diseases, such as autism, dyslexia, or learning difficulties.

Data collection took place in Norfolk and Norwich University Hospitals NHS Foundation Trust, UK between October 2024 and July 2025. CGM device was fitted whilst the consented participants were still in-patients. For further details on the recruitment please consult the methods of our feasibility study [18].

### CGM device and POCT comparison

We used the Dexcom G7 real-time CGM (rt-CGM) device which is licensed for use in children and adults and available for consumers to directly purchase. The kits consist of auto applicator kits with sensors with transmitter and receivers to be used in the blind mode. We provided blinded receivers to all participants and clinical staff which were paired with their sensors in advance.

The sensors were applied to the back of the upper arm. Interstitial glucose readings were measured continuously, and data was transmitted from the sensor every 5 minutes. Time in range (TIR) was established, using a range set between 4 mmol/L to 10 mmol/L in the receiver, which was the default setting, and similar to recommendations [20], [21], [22].

Participants wore the CGM sensors for a maximum of 10 days during their hospital stay. At the same time, POCT measurements were taken a minimum of 4 times a day, as per clinical protocols. If a hypoglycaemic episode was noticed via POCT, additional POCT measurements were taken, pending clinical need. For the analysis we matched the POCT readings closest to the CGM readings. Specifically, we paired POCT measurement with CGM according to the nearest measurement of CGM; ±2 minutes. The difference between the number of POCT (n=895) and paired measurements (n=767) is due to filtering data to 2.2-22.2 mmol/L to avoid potential bias of ceiling effect of measurements and lack of CGM measurements to match due to battery run out (n=2), coming off the sensor earlier than anticipated (n=1), misreading (n=1). Additionally, one paired measurement was excluded due to a clinically implausible discrepancy between readings.

## Statistical Analyses

All statistical analyses were conducted using RStudio (R version 4.3.2; R Foundation for Statistical Computing, Vienna, Austria). Descriptive statistics were used to summarise participant characteristics, clinical data and glucose-related metrics. Continuous variables were reported as mean with standard deviation (SD) or median, while categorical variables were summarised using frequencies and percentages.

Agreement between CGM and POCT measurements was assessed using Bland–Altman analysis. For each paired measurement, the mean of CGM and POCT values and their difference (CGM − POCT) were calculated.

Bias was defined as the mean difference, and 95% limits of agreement (reported as upper and lower LoA) were calculated as the bias ±1.96 standard deviations (SD).

To evaluate clinical accuracy, we conducted a Clarke Error Grid (CEG) analysis, classifying paired CGM–POCT values into Zones A–E according to their potential clinical impact. The proportion of measurements within the clinically acceptable zones (A and B) was reported. Analytical accuracy was further quantified using the mean absolute relative difference (MARD), median absolute relative difference (median ARD), correlation analysis (R^2^) and ±20%/1.1 mmol/L which represents the proportion of CGM values within ±20% of POCT readings >5.6 mmol/L or within ±1.1mmol/L of POCT values ≤5.6 mmol/L, and expressed as a percentage.

## Ethics

This study has been reviewed and approved by Health Research Authority (HRA) and Health and Care Research Wales (HCRW) on 23^rd^ August 2024 (IRAS project ID: 344263, REC reference: 24/LO/0441).

## Results

### Participants

Thirty-two participants were recruited, and with 30 participants included in the analyses. Two participants were excluded: one due to more than 50% missing CGM readings during the 10-day study period; and the other because they were not taking antidiabetic medication.

The participant characteristics are given in Table 1.

**Table 1.** Participant characteristics.

| Participant characteristics | Mean | Median | Min | Max |
| --- | --- | --- | --- | --- |
| Age (years) ± SD | 78.7 ± 6.7 | 78 | 67 | 94 |
| BMI (kg/cm<sup>2</sup>) ± SD | 31.2 ± 10.9 | 28.3 | 20.7 | 74.3 |
| HbA1c (mmol/mol) ± SD | 75.1 ± 29.5 | 67 | 45 | 154 |
| Medications (N) | 10.5 ± 4.4 | 11 | 3 | 20 |
| Hospitalisations (N) in the last 12 months (excluding the time of recruitment) | 0.86 ± 1.5 | 1 | 0 | 8 |
| Length of T2DM (Years) | 14.9 ± 6.4 | 12.5 | 0.6 | 38 |

Twenty-four (80%) participants had at least 4 long-term conditions at admission. Only 1 participant (3.3%) had a previous diagnosis of dementia which was Lewy body dementia.

Nine (30%) participants were admitted to the hospital due to diabetes (diabetic ketoacidosis; DKA, hypoglycemia, and hyperglycaemic hyperosmolar state; HHS), whilst 1 participant (3.3%) was at hospital due to cognitive impairment and confusion. Ten (33.3%) participants were at hospital due to a fall accompanying both diabetes and cognition related reasons.

At the 10 day follow up assessment, sixteen participants (53.3%) were still inpatients, including two (6.6%) at different hospitals for rehabilitation. Two participants (6.6%) were discharged to a care home or nursing home, while the other fourteen participants (46.6%) were at home by the time the visit was scheduled.

### Medications

Most of the participants were treated with insulin; solely or combined therapy, while 43.4% were only on oral glucose anti-diabetes agents, with 33.3% using both insulin and oral agents (Table-2). Due to diabetic foot disease or foot ulcers, 56.6% were regularly using topical agents. Single nutrient supplements (mostly vitamin D, folic acid and thiamine) and multivitamins were commonly used (73.3%).

**Table 2.** Medication classes.

| **Anti-Diabetic Medications\*,\*\*** | **Total Number of participants N (%)** |
| --- | --- |
| **Insulin (solely)** | 7 (23.3%) |
| **Insulin& oral antidiabetics** | 10 (33.3%) |
| **Metformin** | 16 (53.3%) |
| **SGLT-2 inhibitors** | 9 (30%) |
| **DPP4 inhibitors** | 4 (13.3%) |
| **Sulphonylurea** | 3 (10%) |
| **GLP-1 receptor agonists** | 1 (3.3%) |
| **Commonly prescribed other medications\*,\*\*** |  |
| **Statins** | 20 (66.6%) |
| **Anticoagulants/Antiplatelets** | 20 (66.6%) |
| **Topical agents** | 17 (56.6%) |
| **Analgesics** | 15 (50%) |
| **Antihypertensives\*\*\*** | 15 (50%) |
| **Gastric acid suppressive therapy\*\*\*\*** | 11 (39%) |
| **Antibiotics** | 8 (26.6%) |

*\*The medications are the regular medications of our participants recorded at admission.*
*\*\*There was overlap between the groups; for example, some metformin users were also prescribed DPP4 inhibitors, and some inpatients receiving statins were also taking anticoagulants.*
*\*\*\* Antihypertensives include any ACE inhibitors/ARBs, beta blockers, calcium channel blockers, thiazide diuretics and alpha blockers.*
*\*\*\*\*Acid suppressive therapy defines proton pump inhibitors and H2 receptor antagonists.*

### CGM and POCT: TIR, TAR, TBR

Fourteen (46.6%) participants wore their CGM continuously in hospital, with CGM active for a mean of 89.9% of the time (range 86% - 95%).

CGM-derived mean glucose time in range (TIR= 4-10 mmol/L) was 36.2% (range 0-90%), time above range (TAR ≥ 10 mmol/L) was 62.7% and time below range (TBR ≤ 3.9 mmol/L) was 1.03%.

Overall, CGM data were very similar and consistent to POCT glucose measurements (p=0.97) (Table 2). However, there were individual differences between CGM and POCT, with some individuals showing no TBR (0%) via POCT, whilst CGM observed results 3% and 8% TBR within the same participants.

**Table 2.**
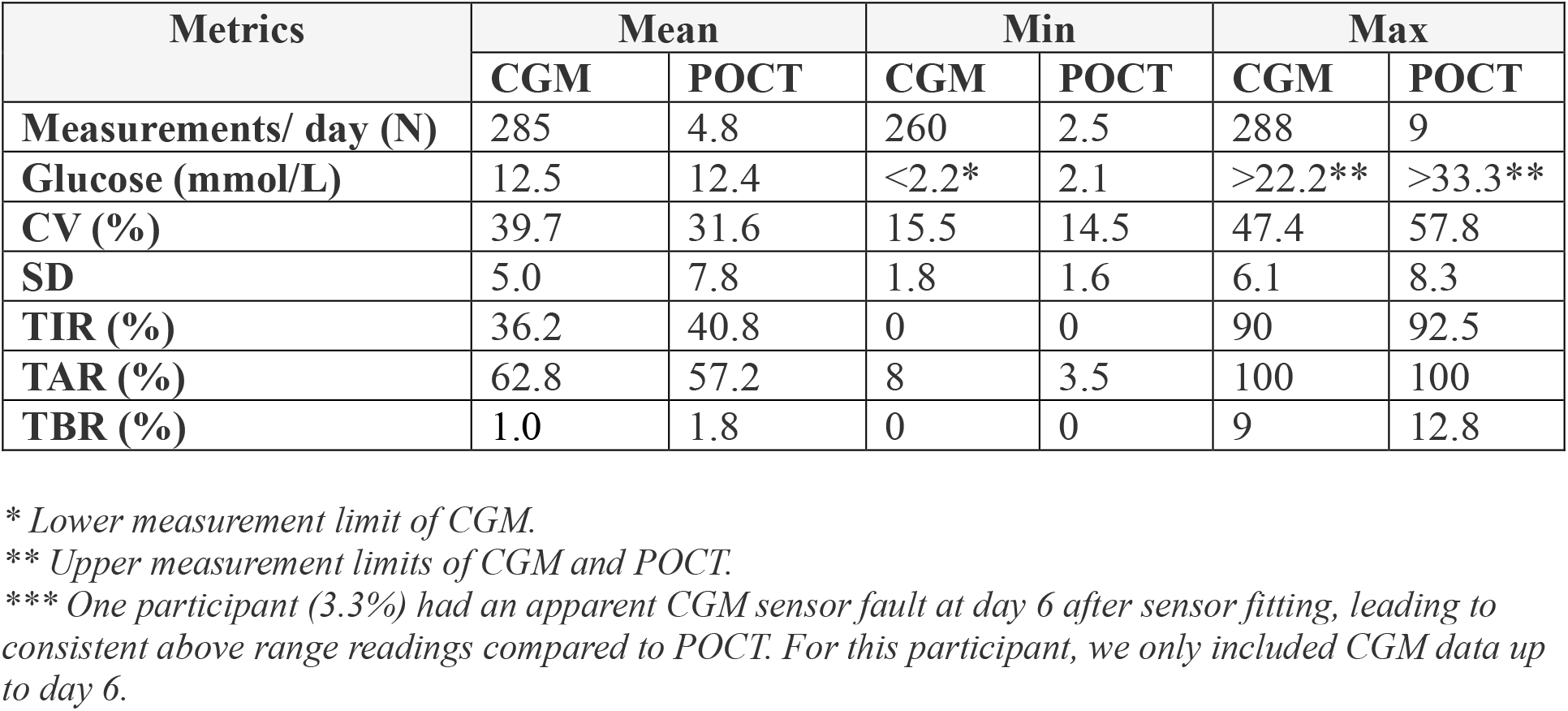
CGM and POCT metrics.

### CGM and POCT: MARD, Bias, Clinical Risk Assessment

767 matched CGM readings and POCT reference values were evaluated using a filtered scatter plot (Figure 1). The total number of CGM and POCT readings, along with available time-matched CGM and POCT readings included in the accuracy analyses are shown Figure 2 (Appendix). Overall, the CGM and POCT measurements demonstrated a strong correlation (R^2^=0.82). A systematic mean bias of 0.88 mmol/L was observed, indicating that the CGM sensor tended to overestimate glucose levels relative to the POCT reference. The overall MARD was 17.43% consistent with an acceptable level of numerical accuracy for devices in this category. In addition, 72.3% of paired readings fell within ±20% or ±1.1 mmol/L of the reference value. Additional metrics describing CGM accuracy and reliability are given in Table 3.

**Table 3.** Summary of CGM accuracy and reliability metrics.

| Metrics | Filtered Data (2.2-22.2 mmol/L) |
| --- | --- |
| MARD (%) | 17.43 |
| Median ARD (%) | 12.24 |
| $R^2$ (Correlation) | 0.82 |
| $\pm 20\%/1.1$ mmol/L (%) | 72.32 |
| Mean bias ±SD | 0.88 ± 2.15 mmol/L |
| Upper Limit of Agreement (95%) | +5.09 mmol/L |
| Lower Limit of Agreement (95%) | -3.33 mmol/L |

**Figure 1.**
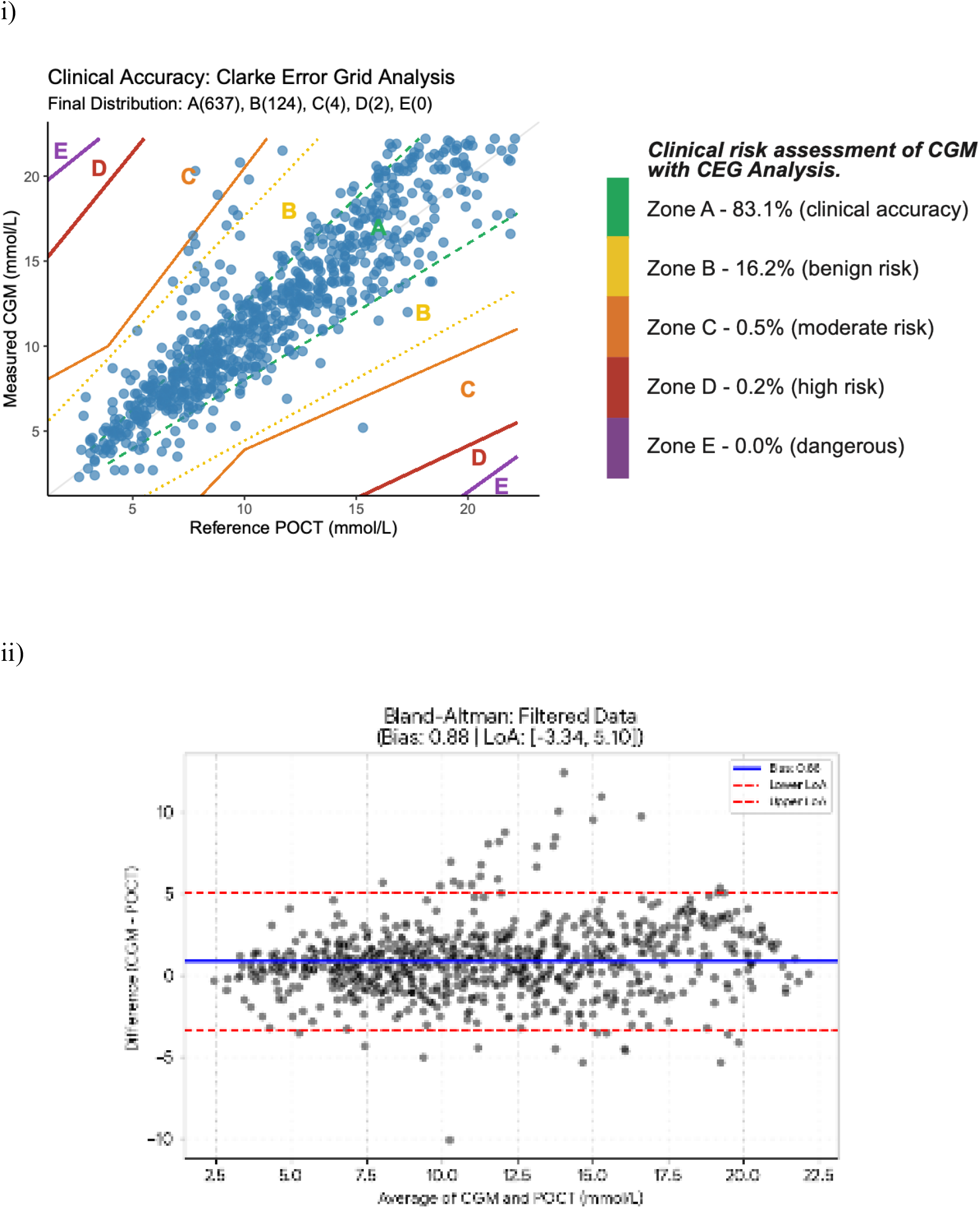
Clinical risk assessment of CGM with CEG and Bland-Altman Analysis. i) Scatter plot diagram showing clinical accuracy of blood glucose levels measured by CGM according to POCT reference values within the filtered range (2.2 – 22.2 mmol/L). ii) Scatter plot diagram demonstrating bias (0.88), lower LoA (-3.33 mmol) and upper LoA (+5.09 mmol/l) of CGM compared to POCT as the reference.

**Figure 2.**
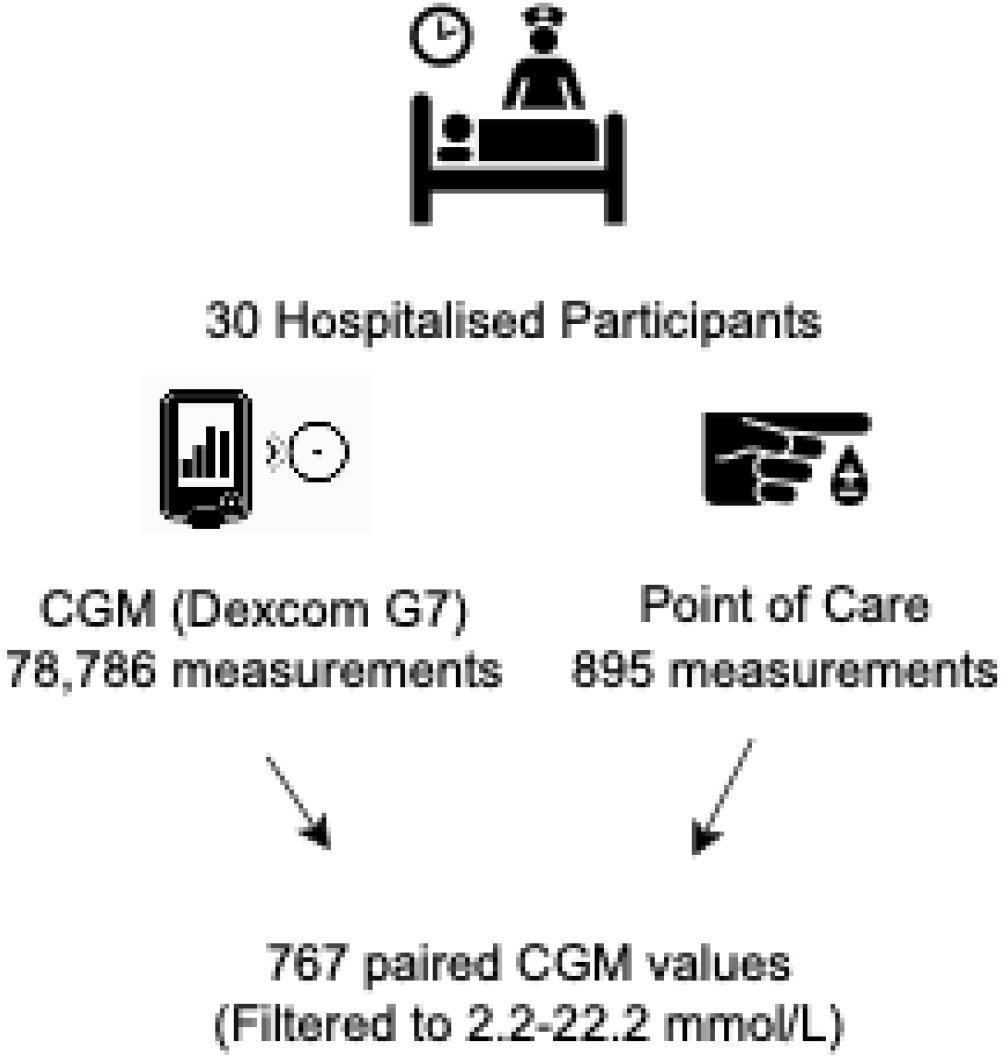
Summary of time-matched CGM and POCT readings used for accuracy analyses. Figure showing the number of CGM and POCT measurements and paired values according to the nearest measurement of CGM; ±2 minutes. The difference between the number of POCT and paired measurements is due to 1. filtering data to 2.2-22.2 mmol/L to avoid potential bias of ceiling effect of measurements 2. lack of CGM measurements to match.

CEG analysis revealed that 99.3% of the data points fell within the clinically acceptable zones (Zone A: 83.1% and Zone B: 16.2%) (Figure 1). These findings confirm the clinical safety of the device, as the vast majority of glucose readings would lead to correct or benign treatment decisions.

### Hypoglycaemia

Nine participants (30%) experienced CGM-detected hypoglycaemia (≤ 3.9mmol/L) during. While four participants experienced only one hypoglycaemic CGM event, others had multiple hypoglycaemic CGM events (median=2.5, max=8, min=1).

One participant was discharged at the time they experienced hypoglycaemia; therefore, POCT-derived glucose measurements were unavailable for comparison with their CGM readings. For the remaining 8 participants who had CGM-derived hypoglycaemic event, we assessed the extent to which these events were captured by POCT during hospital monitoring. Across thes participants, CGM identified 27 hypoglycaemic events, of which 17 (63%) were not detected by POCT hypoglycaemic events (Table 5). With the exception of a single event (ID20, event 4), all POCT-confirmed hypoglycaemic events were recorded with a delay relative to CGM, ranging from at least 34 minutes to as long as 4 hours (ID20, event 5) after the initial low CGM reading. All 8 participants experiencing hypoglycaemia (n=8) had level 2 hypoglycaemic readings (< 3.0 mmol/L), with only one having a level 1 hypoglycaemic event. Table 6 (Appendix) shows more details in mean glucose per hypoglycaemic event, time spent in hypoglycaemia, POCT-capture status, and the time interval between first CGM and POCT readings for each participant.

**Table 5.** Hypoglycaemic Events by CGM and POCT Capture Rate.

| **Participants** | **Number of events by CGM** | **Events missed by POCT N (%)** |
| --- | --- | --- |
| **ID02** | 1 | 1 (100%) |
| **ID03** | 7 | 4 (57.1%) |
| **ID13** | 4 | 2 (50%) |
| **ID14** | 1 | 1 (100%) |
| **ID20** | 8 | 4 (50%) |
| **ID21** | 1 | 0 (0%) |
| **ID24\*** | 1 | N/A |
| **ID31** | 3 | 3 (100%) |
| **ID32** | 2 | 2 (100%) |
| **Total** | **27** | **17 (62.9%)** |

*\*ID24 was discharged by the time they experienced hypoglycaemia. We excluded them from analysis.*

## Discussion

Our findings show that CGM is a safe, clinically acceptable and reliable tool for older inpatients with type 2 diabetes and cognitive impairment, and that it captures more hypoglycaemic episodes than POCT. CGM may therefore complement existing POCT measurements, which are still required to check abnormal readings and guide clinical decision making in in critically ill inpatients [23], [24]. This is the first study to investigate the accuracy of real time CGM in a non-intensive care unit (non-ICU) hospital setting specifically in people with comorbid type 2 diabetes and cognitive impairment.

Overall, CGM-derived glucose metrics showed high consistency with POCT, with no significant group-level differences. However, notable individual-level discrepancies occurred. Some participants showed no TBR with POCT, whilst CGM identified 3-8% TBR for the same individuals. Given the far greater sampling frequency of CGM compared to intermittent POCT (average 4.8 tests per day in our study), it is expected that CGM would detect more dysglycaemic episodes. Our results are similar to previous hospital setting studies [15], although some studies have reported higher TIR with CGM measurements, but similar TBR to POCT [13], [25]. These differences may reflect variation in study design, as previous studies included mixed cohorts of people with type 1 diabetes and type 2 diabetes across a broad age range, while our sample comprised only older adults with type 2 diabetes and cognitive impairment. Furthermore, we also used a newer generation CGM sensor (Dexcom G7), which has a lower reported MARD (8.2%) than older sensors such as the Abbott Freestyle Libre pro (12.3%) and Medtronic Guardian sensor 3 (9.6%) [12], [26]. Together, our TIR, TAR and TBR findings show that CGM and POCT have broadly comparable glycaemic readings in this specific population.

Regarding accuracy, the blinded sensors without real-time calibrations and adjustments produced a MARD of 17.4%, with 99% of paired values falling within zones A and B in Clarke Error Grid. Although many CGM accuracy studies report slightly lower MARD values (11.6%-17.1%) [11], [13], [17], [27], [28], [29], [30], these differences likely reflect broader diabetes and age populations, older sensor generations and inclusion of people with advanced renal dysfunction, who were excluded in our study. Further, some previous studies used intensified POCT measurements (every 5 or 15 minutes), which artificially improves agreement between POCT values and CGM. Notably, one study reported a MARD of 19.2%, and t±20%/1.1 mmol/L accuracy of only 59.5% in a mixed type 1 diabetes and type 2 diabetes cohort, which is lower than our 72.3% [16]. Further comparisons are needed to clarify how diabetes type, age and comorbidities affect differences between CGM and POCT. Despite the differences in MARD differences, our CEG results closely match existing literature (96.5%-99%) [11], [13], [16], [29], supporting the clinical safety of CGM. Previous work reported that CGM accuracy was lower within hospital settings compared to community use, possibly due to reduced opportunities for calibration [16]. Although our sensors were blinded and calibrated only conducted at the beginning of the study, accuracy remained high [12], [31]. Overall, our CEG analysis and ±20%/1.1 mmol/l analyses confirm that CGM is a safe tool for hospital monitoring, though low readings should always be checked by POCT [13], [14], [25], [31].

Our data on hypoglycaemia are particularly noteworthy. POCT missed the majority of CGM-detected hypoglycaemic episodes (62.9%). Some episode were brief or nocturnal, but a significant proportion were prolonged and occurred during the daytime. This is clinically important, since inpatient hypoglycaemia is associated with prolonged length of stay, higher hospital costs, increased readmission rates and higher mortality [31]. Consensus guidelines recommend that older people should spend less than 15 minutes per day in hypoglycaemia range and maintain TBR <1% [32]. Only 11.1% of hypoglycaemic episodes in our cohort lasted less than 15 minutes, and only one participant of those who experienced hypoglycaemia met this guideline threshold. Approximately one third of participants experienced level 1 hypoglycaemia, and many experienced level 2 hypoglycaemias which requires more vigilance and therapeutic adjustment. These findings reinforce the vulnerability of this population and the limitations of POCT.

Our findings support previous evidence that CGM detects earlier, more frequent, and more prolonged hypoglycaemia, including nocturnal events, compared with POCT [13], [14], [15], [25]. Combined with RCT evidence showing improved glycaemic levels and reduced hypoglycaemia when using CGM in non-ICU in-patients on insulin therapy, compared to POCT alone [33], [34], our findings highlight the clinical value of CGM in hospitalised older adults with cognitive decline, who are at particularly high risk of unrecognised hypoglycaemia.

Despite the novelty of our findings, several limitations must be acknowledged. First, this was a single-centre observational study in a relatively small sample size. Replication in larger cohorts and across different hospital settings is essential. participants were admitted for a range of acute conditions, which may have influenced both cognitive performance and glycaemic outcomes. We mitigated this by excluding certain health conditions and only recruiting participants with capacity to consent, thereby reducing confounding from delirium or acute cognitive decline. Third, direct comparison between infrequent POCT results and high frequency CGM data is challenging. Some guidelines recommend assessing CGM accuracy using high-precision laboratory measurement [27], [32], [35], [36], but this was not feasible within our observational, care-as-usual study design. Nevertheless, our findings mirror previous studies and reflect real-world hospital practice, supporting their validity.

In conclusion, CGM as a safe, reliable and clinically informative monitoring tool for older inpatients with type 2 diabetes and cognitive impairment and detects significantly more hypoglycaemia than conventional POCT. However, dysglycaemic readings, particularly hypoglycaemia should always be verified via POCT before clinical intervention. Larger studies across diverse settings are needed to optimise inpatient glucose management pathways and determine the optimal integration of CGM into routine care for this vulnerable population.

## Data Availability

All data produced in the present study are available upon reasonable request to the authors.

## Data Availability

All data produced in the present study are available upon reasonable request to the authors.

## Acknowledgements

We would like to thank the participants in this study for devoting their time for this research. The views expressed are those of the author(s) and not necessarily those of the NIHR or the Department for Health and Social Care.

## Appendix

**Table 6.** Time Spent in Hypoglycaemia and Time-of-Day Distribution of CGM Events and POCT Capture.

| Partici<br>pants | The<br>number<br>of<br>hypoglyc<br>aemic<br>events | Mean<br>gluco<br>se<br>(mmo<br>l/L) | Min<br>gluco<br>se<br>(mmo<br>l/L) | Max<br>gluco<br>se<br>(mmo<br>l/L) | Time<br>spent in<br>hypoglyc<br>aemia to<br>CGM<br>(~minute<br>s) | Capture<br>d by<br>POCT | If capture<br>d, time<br>gap<br>between<br>n CGM<br>and<br>POCT<br>(~minu<br>tes<br>after) | Noctu<br>rnal<br>(12am<br>- 6am) |
| --- | --- | --- | --- | --- | --- | --- | --- | --- |
| **ID02** | 1 | 3.6* | 3.2 | 4 | 115 | No | N/A | Yes |
| **ID03** | 1 | 3.6 | 3.6 | 3.8 | 50 | Yes | 41 | No |
|  | 2 | 3.4 | 3.3 | 3.8 | 75 | Yes | 111 | No |
|  | 3 | 3.2 | 2.8 | 3.8 | 70 | No | N/A | No |
|  | 4 | 3.6 | 3.4 | 3.9 | 30 | Yes | 34 | No |
|  | 5 | 3.3*,*<br>* | 2.2 | 4.4 | 130 | No | N/A | No |
|  | 6 | 3.2* | 2.7 | 4.3 | 50 | No | N/A | No |
|  | 7 | 3.3* | 2.2 | 4.4 | 105 | No | N/A | No |
| **ID13** | 1 | 3.9* | 3.8 | 4.1 | 40 | No | N/A | Yes |
|  | 2 | 2.9** | 2.2 | 3.9 | 105 | Yes | 45 | No |
|  | 3 | 3.8* | 3.5 | 4.5 | 95 | No | N/A | No |
|  | 4 | 3.4* | 2.7 | 5.4 | 85 | Yes | 61 | No |
| **ID14** | 1 | 3.4 | 2.8 | 3.9 | 15 | No | N/A | No |
| **ID20** | 1 | 3.6 | 3.5 | 3.8 | 130 | No | N/A | Yes |
|  | 2 | 3.0 | 2.6 | 3.6 | 245 | No | N/A | Yes |
|  | 3 | 3.5 | 3.2 | 3.8 | 130 | Yes | 85 | No |
|  | 4 | 3.8 | 3.8 | 3.8 | 5*** | Yes | 8<br>(before<br>****) | No |
|  | 5 | 3.2 | 2.7 | 3.8 | 285 | Yes | 267 | Yes |
|  | 6 | 3.3 | 2.8 | 3.8 | 290 | No | N/A | Yes |
|  | 7 | 3.8 | 3.8 | 3.8 | 10*** | No | N/A | Yes |
|  | 8 | 3.5 | 3.2 | 3.8 | 65 | Yes | 40 | No |
| **ID21** | 1 | 3.8* | 3.5 | 4.2 | 85 | Yes | 72 | Yes |
| **ID24\*\***<br>**\*\*\*** | 1 | 3.0** | 2.2 | 3.9 | 10*** | N/A | N/A | No |
| **ID31** | 1 | 3.4* | 2.4 | 4.8 | 205 | No | N/A | No |
|  | 2 | 3.3 | 3.4 | 3.9 | 40 | No | N/A | No |
|  | 3 | 2.7*,*<br>* | 2.2 | 4.1 | 230 | No | N/A | Yes |
| **ID32** | 1 | 2.9*,*<br>* | 2.2 | 4.1 | 525 | No | N/A | Yes |
|  | 2 | 2.6** | 2.2 | 3.8 | 520 | No | N/A | Yes |
| **Total** | **28** | **3.4** |  |  | **≈149 (all)**<br>**≈74.7 (diurnal)** | **Capture**<br>**d=10,**<br>**Missed=17**<br>**Nocturnal**<br>**missed=9** | **≈84 (all)**<br>**≈59.5 (diurnal)** | **Yes=11,**<br>**No=17** |

*\*Participant had >3.9mmol/L readings during hypoglycaemic event, however, did not exceed 20 minutes (2 readings) according to CGM.*
*\*\*Participant had <2.2mmol/L readings which are lower cut off CGM. We included them into our statistics as 2.2mmol/L to have valid data.*
*\*\*\*These hypoglycaemic results may not be defined as hypoglycaemic events due to uncertainty of time spent at hypoglycaemia. Events are required to last at least 15 minutes [37]. Therefore, we excluded these three results from the average value (≈149).*
*\*\*\*\*Participant's (ID20) 4<sup>th</sup> hypoglycaemic event was captured 8 minutes before CGM showed low reading. Except this event, other events were captured after CGM showed low glucose results.*
*\*\*\*\*\*We only included low readings if POCT is available for the event to match CGM results. ID24 did not have hospital POCT records the time they experienced hypoglycaemia.*

